# Making a Case for an Autism-Specific Multimorbidity Index: A Comparative Cohort Study

**DOI:** 10.1101/2024.02.23.24303273

**Authors:** Filip Sosenko, Dewy Nijhof, Laura McKernan Ward, Deborah Cairns, Laura Hughes, Ewelina Rydzeswka, CVD-COVID-UK/COVID-IMPACT Consortium

**Affiliations:** University of Glasgow, School of Mental Health and Wellbeing; University of Dundee, Health Informatics Centre; University of Edinburgh, School of Health in Social Science

**Keywords:** Autism Spectrum Disorder, Multimorbidity, Multimorbidity Index, COVID-19, Mortality, Long-term Conditions

## Abstract

Autistic people experience challenges in healthcare, including disparities in health outcomes and multimorbidity patterns distinct from the general population. This study investigated the efficacy of existing multimorbidity indices in predicting COVID-19 mortality among autistic adults and proposes a bespoke index, the ASD-MI, tailored to their specific health profile. Using data from the CVD-COVID-UK/COVID-IMPACT Consortium, encompassing England’s entire population, we identified 1,027 autistic adults hospitalized for COVID-19, among whom 62 died due to the virus. Employing logistic regression with 5-fold cross-validation, we selected diabetes, coronary heart disease, and thyroid disorders as predictors for the ASD-MI, outperforming the Quan Index, a general population-based measure, with an AUC of 0.872 versus 0.828, respectively. Notably, the ASD-MI exhibited better model fit (pseudo-R2 0.25) compared to the Quan Index (pseudo-R2 0.20). These findings underscore the need for tailored indices in predicting mortality risks among autistic individuals. However, caution is warranted in interpreting results, given the limited understanding of morbidity burden in this population. Further research is needed to refine autism-specific indices and elucidate the complex interplay between long-term conditions and mortality risk, informing targeted interventions to address health disparities in autistic adults. This study highlights the importance of developing healthcare tools tailored to the unique needs of neurodivergent populations to improve health outcomes and reduce disparities.

## Introduction

Autism Spectrum Disorder (ASD) is a neurodevelopmental disorder characterised mainly by differences in social communication and interaction, and repetitive and restrictive behaviours, including differences in sensory preferences (1). Autistic people are estimated to account for 0.6 to 1.0% of the global population (2–4). They experience substantial health inequalities, including multimorbidity (5,6), defined as the presence of at least two long-term conditions (LTCs) (7). ASD has been proposed as a potential risk factor for the development of COVID-19 infections due to a number of factors including; experiencing difficulties in maintaining social distancing (8), and use of atypical antipsychotics, such as risperidone, frequently prescribed to individuals with ASD (9), which has been shown to disrupt immune responses (10), and therefore increase the risk of COVID-19 infection and mortality.

Autistic people have a different pattern of multimorbidity compared to the general population (11,12) which can result in premature mortality (13,14). What is more, literature suggests that autistic people are likely to experience poor physical health (12,15), high prevalence of multimorbidity (16), and premature mortality (13,17,18). This highlights the necessity of capturing the full burden of multimorbidity to detect poor health and provide suitable care.

Multimorbidity indices, which are used to predict the prognosis of patients based on their medical history or to measure the comorbidity burden, are currently based on the general population (19–21), and may not be applicable to the population with ASD. Multimorbidity indices use previous and current diagnoses as predictors for the outcome of interest, such as quality of life, hospital admissions, healthcare use, and mortality (22,23). Alternatively, the index is used to control for long-term conditions or multimorbidity while studying other associations.

A variety of multimorbidity indices exist, and characteristics amongst these instruments vary in terms of included LTCs, weighting of LTCs and even outcome measures (22). The most used and well-known index is the Charlson Comorbidity Index (CCI) which uses scores for 17 LTCs to predict the ten-year risk of death within one year after hospital admission (19). Since its conception, several modifications of the CCI have been created, ranging in which LTCs are included, total number of diseases included, weights of individual LTCs and overall score construction (22). The most well-known modifications include the Elixhauser Index (including 30 – or, for some variants, 31 – LTCs) (20) and the van Walraven (VW) variant of the Elixhauser Index, which includes a weighted summary score for streamlined use, based on the 30 LTCs from the Elixhauser Index (21). The Quan Index is a relatively recent mortality prediction tool utilized in healthcare settings, primarily focused on in-hospital mortality risk assessment. Unlike the Charlson Index, which includes 17 Long-Term Conditions (LTCs), the Quan Index streamlines the assessment by incorporating only 12 LTCs (24). These LTCs are determined based on International Classification of Diseases, 10th Revision (ICD-10) codes. One notable feature of the Quan Index is its utilization of up-to-date population health data from six different countries to inform the construction of weights assigned to each LTC, enhancing the predictive accuracy of the index (24).

Most indices use scores to estimate prognosis, with higher scores usually being indicative of more severe risk of death. These scores are usually additive, where the presence of multiple LTCs will lead to a higher score. Scores are based on weights for each condition, which usually are derived from the modelling of the risk, in which the weight quantifies their contribution towards the outcome. The long history of development and validation of these instruments has resulted in establishing strong evidence of associations between multimorbidity and mortality risk, decline in physical and mental functioning, and quality of life (22,23). However, less research has focused on the selection criteria for inclusion of LTCs and most indices are constructed for the general population (22).

As common physical and mental LTCs differ in the population with ASD compared from those in the general population (13,25,26), this may be of importance. For example, autistic people have a high burden of mental health conditions such as anxiety, depression and schizophrenia (14,27). Autistic people are also more likely to experience co-occurring physical conditions, such as epilepsy, autoimmune disorders and obesity (14,27). Therefore, the effectiveness and sensitivity of the established multimorbidity indices for the prediction of health outcomes in the autistic population and subsequent treatment based on those predictions warrants an investigation into their validity. Indeed, studies examining the association between LTCs and mortality in autistic people make use of established multimorbidity indices, such as the CCI, but include LTCs known to be prevalent in the autistic population (14,27). These studies found that the included LTCs, such as epilepsy, mental health conditions (e.g., bipolar disorder, schizophrenia, major depressive disorder) or intellectual disabilities carried higher risk of death in the autistic population (14,27). This suggests that traditional indices may not accurately reflect the health profile or capture the full extent of multimorbidity and its concurrent risk for the autistic population.

To our knowledge, there has been no previous research into the effectiveness of using existing multimorbidity indices to investigate health outcomes in autistic people nor does a multimorbidity index specifically tailored for autistic populations exist. As such, the current study aims to address this gap in the existing literature by investigating the effectiveness and sensitivity of an established multimorbidity index compared to a specifically tailored index, hereafter called Autism Spectrum Disorder Multimorbidity Index (ASD-MI), in predicting COVID-19 mortality within the autistic population. This study represents a supplementary analysis derived from a broader investigation examining COVID-19 outcomes in autistic adults (28,29). As such, the study design employed a convenience sampling approach, necessitated by the availability of relevant data sources.

## Methods

### Data sources

This was part of a larger cross-sectional study using a whole-country population covering England (30,31). It was conducted on behalf of, and accessed data made available in NHS England’s Secure Data Environment service for England, the CVD-COVID-UK/COVID-IMPACTConsortium (coordinated by the BHF Data Science Centre). The datasets used for the current study were Hospital Episode Statistics (HES), COVID-19 Hospitalisation in England Surveillance System (CHESS), Second Generation Surveillance System (SGSS), General Practice Extraction Service (GPES) Data for Pandemic Planning and Research (GDPPR) and Civil Registry Deaths from the Office for National Statistics (ONS-D). All but the last of these datasets are updated through the National Health Service (NHS) and processed by NHS England who then opens up the data for secondary use. The ONS-D dataset is updated through the ONS. Data from 1 March 2020 up to and including 31 December 2021 was used. Data linkage was conducted using an anonymised version of the NHS number (a ten-digit number used as a unique identifier within the UK NHS to identify patients). Ethical approval was obtained from the CVD-COVID-UK/COVID-IMPACT Consortium and the University of Glasgow Ethics Committee.

Autistic people were identified using SNOMED concepts sourced from the GDPPR and HES ‘Autism Diagnosis Codes’ cluster^1^ (cluster AUTISM COD version 2021/12/21). With regards to the long-term conditions considered as ‘candidates’ for the ASD-MI, thirty-five long-term conditions common to the UK population were identified (32) (see Supplementary materials, table 1). One-year look-back window (in HES) was used to identify patients with a given condition, counting from the date of COVID-19 admission.

Demographic (age, sex, ethnicity, Index of Multiple Deprivation (IMD)) and LTCs were explored using percentages of total autistic population. IMD decile is a measure used in the United Kingdom to assess relative deprivation across different areas or regions. The IMD ranks from 1 (most deprived) to 10 (least deprived) based on several indicators of deprivation, such as income, employment, education, health, crime, and living environment. In our analyses, instead of having 10 distinct deciles, the IMD rankings were grouped into 5 categories, each representing a range of deprivation levels. This grouping simplifies the interpretation of deprivation levels and facilitates comparisons between areas.

The decision to compare our ASD-MI to the Quan Index rather than the Charlson Index was carefully considered. Firstly, our study was constrained by the requirement to use ICD-10 code mapping, which is a feature supported by the Quan Index but not the Charlson Index (24). Furthermore, the Quan Index’s more recent update and incorporation of contemporary population health data from six countries provided a distinct advantage (24). This up-to-date information enhances the predictive capabilities of the Quan Index, aligning with our study’s objective of accurately predicting in-hospital mortality risk among autistic adults with COVID-19. To predict in-hospital mortality, the Quan Index includes 12 LTCs instead of the original 17 used in the Charlson Index to construct weights (33) (see Supplementary materials, table 2). Again, a one-year look-back window in HES was used to calculate the patient’s Quan Index score.

### Population

The study sample was drawn from a larger study sample that covered the whole population of England who were alive on 1 January 2020 (31). Adults who had a primary care diagnosis of autism and who were hospitalised for COVID-19 (defined as any CHESS record, or a HES record with an ICD-10 code of U07.1 or U07.2 recorded in any position on the admission diagnosis). ‘COVID-19 cause of death’ was defined as a death that occurred within 28 days from the first day of admission and where COVID-19 was mentioned on the death certificate (either primary or other contributing causes using ICD-10 code U07.1 or U07.2). For the larger study, the total sample with confirmed COVID-19 was 32,372 autistic adults (see Table 1). The sample used to construct the multimorbidity index consisted of 1,027 autistic adults hospitalised for COVID-19, of which 62 died due to COVID-19. Data up to 31 December 2021 was included.

**Table 1.** Demographic and health characteristics of autistic adults hospitalised for COVID-19.

|  | Covid-19 hospitalised adults who subsequently died from covid | Covid-19 hospitalised adults who subsequently did not die from covid | All covid-19 hospitalised adults |
| --- | --- | --- | --- |
| Total N (rounded) | 60 | 965 | 1,030 |

| Demographic information |  |  |  |
| --- | --- | --- | --- |
| Age (median, IQR) | 63.4 [48.9, 75.2] | 31.1 [23.6, 50.0] | 32.3 [24.0, 53.0] |
| Female (%) | 19.4 | 40.6 | 39.3 |
| IMD decile (mean) | 4.8 | 4.6 | 4.6 |
| Ethnicity (%) |  |  |  |
| White | 96.8 | 89.5 | 90.0 |
| Asian | <10 CASES | 3.2 | 3.1 |
| Black | <10 CASES | 2.7 | 2.5 |
| Mixed | <10 CASES | 2.7 | 2.6 |
| Other | <10 CASES | 1.5 | 1.4 |
| Long-term conditions (%) |  |  |  |
| Count of LTC (mean) | 11.0 | 7.4 | 7.6 |
| Alcohol problems | 33.9 | 31.3 | 31.5 |
| Anorexia or bulimia | <10 CASES | 8.3 | 8.1 |
| Anxiety & other neurotic | 64.5 | 58.8 | 59.1 |
| Asthma (currently treated) | 27.4 | 27.9 | 27.9 |
| Atrial fibrillation | 33.9 | 10.4 | 11.8 |
| Blindness and low vision | <10 CASES | 6.7 | 7.0 |
| Bronchiectasis | <10 CASES | <10 CASES | 1.0 |
| Cancer - [New]Diagnosis in last five years | 40.3 | 18.0 | 19.4 |
| Coronary heart disease | 58.1 | 19.6 | 21.9 |
| Chronic kidney disease | <10 CASES | 1.2 | 1.2 |
| Chronic Liver Disease and Viral Hepatitis | 43.6 | 32.5 | 33.2 |
| COPD | 24.2 | 10.0 | 10.8 |
| Dementia | 51.6 | 52.5 | 52.5 |
| Depression | 38.7 | 43.0 | 42.8 |
| Diabetes | 85.5 | 37.8 | 40.7 |
| Diverticular disease of intestine | 43.6 | 25.6 | 26.7 |
| Epilepsy (currently treated) | 43.6 | 21.6 | 22.9 |
| Heart failure | 61.3 | 22.7 | 25.0 |
| Hearing loss | <10 CASES | 6.1 | 6.6 |
| Hypertension | 66.1 | 24.6 | 27.1 |
| Inflammatory bowel disease | 17.7 | 14.6 | 14.8 |
| Irritable bowel syndrome | <10 CASES | 9.3 | 9.4 |
| Migraine | <10 CASES | 5.5 | 5.7 |
| Multiple sclerosis | <10 CASES | <10 CASES | <10 CASES |
| Peptic Ulcer Disease | <10 CASES | 10.8 | 10.9 |
| Parkinson's disease | <10 CASES | 2.1 | 2.3 |
| Prostate disorders | <10 CASES | 6.8 | 7.2 |
| Psychoactive substance misuse (not alcohol) | 64.5 | 62.3 | 62.4 |
| Psoriasis or eczema | <10 CASES | 7.2 | 7.2 |
| Peripheral vascular disease | 22.6 | 7.2 | 8.1 |
| Rheumatoid arthritis | 22.6 | 15.2 | 15.7 |
| Schizophrenia (and related non-organic psychosis) or bipolar disorder | 83.9 | 78.9 | 79.2 |
| Chronic sinusitis | <10 CASES | 1.4 | 1.4 |
| Stroke & transient ischaemic attack | 62.9 | 51.1 | 51.8 |
| Thyroid disorders | 17.7 | 5.6 | 6.3 |

### Data analysis

All data analysis was conducted in Python 3.7 and SQL. The analysis was performed according to a pre-specified analysis plan published on GitHub, along with the phenotyping and analysis code (https://github.com/BHFDSC/CCU030_03).

In the first stage of data analysis, a forward variable selection procedure using logistic regression with 5-fold cross-validation was used to identify which of the 35 long-term conditions (see Supplementary materials, table 3) should be selected for the ASD-MI. Age was included as an additional predictor, as per procedure in the development of the CCI (19). The absence or presence of COVID-19 death was the outcome. Area Under the Curve (AUC) score was used as the model evaluation metric. Predictor selection was stopped when the AUC value of the model could not be further improved by at least 0.001. Once a final model was arrived at, coefficients of the long-term conditions from that model were then used to construct an ASD-MI Score for each condition.

Once the ASD-MI was ready, the second stage of data analysis commenced by assessing predictive performance of a model with the Quan Index and a model with the ASD-MI through 5-fold cross validation with AUC as evaluation metric. The modelling employed logistic regression with the absence or presence of COVID-19 death as the outcome. Age was again included as a control variable.

## Results

### Demographics

The results presented in Table 1 provide a comprehensive overview of the demographic and health characteristics of autistic adults hospitalised for COVID-19, categorised based on subsequent COVID-19 mortality outcomes. Among the total cohort of 1,030 hospitalised adults, 60 individuals died due to COVID-19, while 965 individuals did not. The median age of those who died from COVID-19 was notably higher at 63.4 years (interquartile range: 48.9 to 75.2) compared to survivors, with a median age of 31.1 years (interquartile range: 23.6 to 50.0). A higher proportion of females were observed among survivors (40.6%) compared to those who died from COVID-19 (19.4%). Additionally, the mean IMD decile was similar between both groups, indicating comparable levels of deprivation. Regarding LTCs, individuals who subsequently died from COVID-19 had a higher mean count of LTCs (11.0) compared to survivors (7.4). Furthermore, several LTCs demonstrated notable differences in prevalence between the two groups, with higher proportions observed among individuals who died from COVID-19, although these results should be interpreted with caution due to small numbers.

### Predictor Selection for the ASD-MI

By employing a forward variable selection procedure using logistic regression with 5-fold cross-validation, we systematically evaluated the predictive capabilities of 35 long-term conditions (LTCs) in relation to COVID-19 mortality risk among autistic adults. This approach allowed us to identify the most influential LTCs associated with the outcome of interest. By iteratively assessing the model’s performance using the Area Under the Curve (AUC) score as the evaluation metric, we identified three LTCs that consistently demonstrated the highest predictive power. The selected variables for the ASD-MI were diabetes, coronary heart disease and thyroid disorders. These LTCs emerged as significant contributors to the ASD-MI, reflecting their strong association with COVID-19 mortality risk in this population. This process ensured that the selected LTCs were robust indicators of mortality risk, disregarding indicators that did not add to the predictive power of the model. Table 2 shows the coefficients of the LTCs included in the ASD-MI and their corresponding Odds Ratios. Using the coefficients from the model, the formula for the Multimorbidity Index (MI) was:

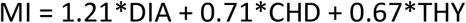

Where DIA refers to diabetes, CHD to coronary heart disease, and THY to thyroid disorders. Each are multiplied by their respective coefficients from the model.

**Table 2.** Logistic Regression results for the models using the Quan Index and the ASD-MI.

|  | Odds Ratio | 95% Confidence Interval | P-value |
| --- | --- | --- | --- |
| **Model with Quan Index** |  |  |  |
| Age at COVID | 1.06 | 1.05, 1.08 | 0.000 |
| Quan Index score | 1.16 | 0.98, 1.37 | 0.081 |
| Constant | 0.00 | 0.00, 0.01 | 0.000 |

| **Model with ASD-MI** |  |  |  |
| --- | --- | --- | --- |
| Age (at COVID) | 1.05 | 1.03, 1.07 | 0.000 |
| Multimorbidity Index score | 2.72 | 1.78, 4.14 | 0.000 |
| Constant | 0.00 | 0.00, 0.01 | 0.000 |

### Comparative Predictive Performance of the two models

The logistic model using the ASD-MI had a higher AUC value of 0.872 than the model using the Quan Index, which had an AUC value of 0.828. Table 2 shows the regression results for both models using the Quan Index and the ASD-MI. Further, model fit is better for the model using the ASD-MI (pseudo-R^2^ 0.25) than for the model using Quan Index (pseudo-R^2^ 0.20).

## Discussion

This feasibility study aimed to investigate whether there is a need for a specially tailored multimorbidity index to predict COVID-19-related mortality in autistic adults. Using LTCs that more accurately describe the health profile of the autistic population, the results show that our ASD-MI is moderately better at predicting the risk of COVID-19 death amongst autistic adults than a general population-based index. This study is a case in point to show the potential for development for general poor health outcomes specific to autistic adults, especially if trained for mortality outside of a COVID-19 context.

While our results suggest a better prediction of COVID-19 mortality in autistic adults using the three predictors of diabetes, coronary heart disease and thyroid dysfunction, caution should be used when interpreting these results. We emphasise that further research is needed to accurately predict all-cause mortality due to the known associations of these three conditions with COVID-19 (34–36). Current evidence on all-cause mortality in autistic individuals points mainly to external causes, such as suicide, and neurological disorders (13,14) It could well be that the current study’s formulation of the ASD-MI does not generalise well to all-cause mortality in autistic adults. Furthermore, relatively little is known about the morbidity burden of coronary heart disease and thyroid disorders in autistic individuals. This further emphasises the importance of studying the general health profile of autistic individuals and how a specifically tailored multimorbidity index could help in finding target LTCs for future research.

One aspect of the study that needs to be emphasised is that our ASD-MI, similar to the original CCI and Quan Index, was developed purely for the purpose of prediction. Accordingly, predictive power was the only criterion for selecting one model over another. Therefore, like the creators of earlier indices we did not aim to explain why selected LTCs were better predictors than LTCs that were ruled out. And we do not seek to justify LTC selection on grounds of existing knowledge and risk factors for COVID-19 mortality (see (37) for further discussion).

The current study did find significant differences in the predictive value of our ASD-MI and a standard index, despite limiting the data to COVID-19-related deaths in the peak of the global pandemic in England. Future research expanding this methodology to all-cause mortality using a larger window of time may yield larger sample sizes and more generalisable results. This would arguably help improve predicting mortality for autistic adults in healthcare settings. Further, it would help elucidate the multimorbidity profile of autistic adults by virtue of its selection criteria for included LTCs. Furthermore, these findings and future exploration of all-cause mortality may highlight LTCs and combinations of LTCs that are of most urgency to investigate in order to address increased mortality in autistic adults.

### Strengths and limitations

This study has several strengths. It is the first study to investigate and construct a multimorbidity index specifically aimed at autistic adults. Foremost, the utilization of a whole-country population encompassing both autistic individuals and the general population ensures a comprehensive and representative sample, effectively mitigating potential sampling biases. Additionally, the study leverages validated datasets, further fortifying the credibility of the results. Furthermore, the inclusion criteria for autism diagnosis are meticulously defined based on clinical diagnoses, incorporating a wide range of older and newer diagnostic codes pertinent to autism. This comprehensive approach enables the study to capture a diverse spectrum of individuals on the autism spectrum, thereby offering a more nuanced and accurate representation of the autistic population in England. Overall, these strengths collectively underscore the study’s capacity to provide valuable insights into the health profiles and multimorbidity patterns among autistic individuals, contributing to a deeper understanding of their healthcare needs.

One potential limitation stems from the sample characteristics. Given the focus on evaluating the multimorbidity index within the context of COVID-19, the sample composition may be skewed, particularly in terms of age and sex distribution. Notably, while older age has been associated with increased COVID-19 mortality risk, the autistic sample in this study tended to be younger due to the higher rates of autism diagnosis in children and young individuals. Moreover, the male predominance in both COVID-19 cases and autism diagnoses introduces further complexity, potentially influencing the observed outcomes. These inherent biases may limit the extrapolation of study findings to broader populations. However, despite these limitations, this study contributes valuable insights into the health profiles of autistic individuals and underscores the importance of developing tailored tools to address their unique healthcare needs, thereby highlighting avenues for further research and exploration in this area.

Another potential limitation is the possible introduction of an inherent bias in the data due to the use of several code mappings to accommodate information coding in the different types of datasets and analysis tools used which originates from data linkage using multiple systems that use different codes within their electronic health records.

Further, while the follow-up time in the CCI and Quan Index is one year after admission, the current study used a follow-up time of 28 days in order to be consistent with the official definition of COVID-19 death.

## Conclusion

This point in case study demonstrated that the use of our ASD-MI for the adult autistic population provides more accurate predictions of COVID-19 mortality in this population than a multimorbidity index based on the general population. It shows that application of the ASD-MI trained on all-cause mortality the autistic population may be more appropriate than the use of multimorbidity indices based on the general population and could be used to elucidate key differences between the health profiles of autistic and non-autistic people.

## Data Availability

The data used in this study are available in NHS England's Secure Data Environment (SDE) service for England, but as restrictions apply they are not publicly available (https://digital.nhs.uk/services/secure-data-environment-service).
The CVD-COVID-UK/COVID-IMPACT programme led by the BHF Data Science Centre (https://bhfdatasciencecentre.org/) received approval to access data in NHS England's SDE service for England from the Independent Group Advising on the Release of Data (IGARD) (https://digital.nhs.uk/about-nhs-digital/corporate-information-and-documents/independent-group-advising-on-the-release-of-data) via an application made in the Data Access Request Service (DARS) Online system (ref. DARS-NIC-381078-Y9C5K) (https://digital.nhs.uk/services/data-access-request-service-dars/dars-products-and-services).
The CVD-COVID-UK/COVID-IMPACT Approvals & Oversight Board (https://bhfdatasciencecentre.org/areas/cvd-covid-uk-covid-impact/) subsequently granted approval to this project to access the data within NHS England's SDE service for England. The de-identified data used in this study were made available to accredited researchers only. Those wishing to gain access to the data should contact in the first instance.

## Declarations

### Ethics approval and consent to participate

The North East – Newcastle and North Tyneside 2 research ethics committee provided ethical approval for the CVD-COVID-UK/COVID-IMPACT research programme (REC No 20/NE/0161) to access, within secure trusted research environments, unconsented, whole-population, de-identified data from electronic health records collected as part of patients’ routine healthcare. The need for informed consent was waived by the North East – Newcastle and North Tyneside 2 research ethics committee. All methods were carried out in accordance with relevant guidelines and regulations.

### Availability of data and materials

The data used in this study are available in NHS England’s Secure Data Environment (SDE) service for England, but as restrictions apply they are not publicly available (https://digital.nhs.uk/services/secure-data-environment-service).

The CVD-COVID-UK/COVID-IMPACT programme led by the BHF Data Science Centre (https://bhfdatasciencecentre.org/) received approval to access data in NHS England’s SDE service for England from the Independent Group Advising on the Release of Data (IGARD) (https://digital.nhs.uk/about-nhs-digital/corporate-information-and-documents/independent-group-advising-on-the-release-of-data) via an application made in the Data Access Request Service (DARS) Online system (ref. DARS-NIC-381078-Y9C5K) (https://digital.nhs.uk/services/data-access-request-service-dars/dars-products-and-services).

The CVD-COVID-UK/COVID-IMPACT Approvals & Oversight Board (https://bhfdatasciencecentre.org/areas/cvd-covid-uk-covid-impact/) subsequently granted approval to this project to access the data within NHS England’s SDE service for England. The de-identified data used in this study were made available to accredited researchers only. Those wishing to gain access to the data should contact in the first instance.

### Funding

The British Heart Foundation Data Science Centre (grant No SP/19/3/34678, awarded to Health Data Research (HDR) UK) funded co-development (with NHS England) of the Secure Data Environment service for England, provision of linked datasets, data access, user software licences, computational usage, and data management and wrangling support, with additional contributions from the HDR UK Data and Connectivity component of the UK Government Chief Scientific Adviser’s National Core Studies programme to coordinate national COVID-19 priority research. Consortium partner organisations funded the time of contributing data analysts, biostatisticians, epidemiologists, and clinicians.

The associated costs of accessing data in NHS England’s Secure Data Environment service for England, for analysts working on this study, were funded by the Data and Connectivity National Core Study, led by Health Data Research UK in partnership with the Office for National Statistics, which is funded by UK Research and Innovation (grant ref: MC_PC_20058).

The Baily Thomas Charitable Fund funded staff (FS) time on this project.

## Acknowledgements

This work is carried out with the support of the BHF Data Science Centre led by HDR UK (BHF Grant no. SP/19/3/34678). This study makes use of de-identified data held in NHS England’s Secure Data Environment service for England and made available via the BHF Data Science Centre’s CVD-COVID-UK/COVID-IMPACT consortium. This work uses data provided by patients and collected by the NHS as part of their care and support. We would also like to acknowledge all data providers who make health relevant data available for research.

The authors would like to thank The Baily Thomas Charitable Fund for funding staff costs on this study.

## Data Availability Statement

For the purpose of open access, the authors have applied a Creative Commons Attribution (CC BY) licence to any Author Accepted Manuscript version arising from this submission.

## Footnotes

1 Primary Care Domain Reference Set Portal. NHS Digital, https://digital.nhs.uk/data-and-information/data-collections-and-data-sets/data-collections/quality-and-outcomes-framework-qof/quality-and-outcome-framework-qof-business-rules/primary-care-domain-reference-set-portal (accessed 8 August 2023).

## Notes

### Competing Interest Statement

The authors have declared no competing interest.

### Author Declarations

The North East-Newcastle and North Tyneside 2 research ethics committee provided ethical approval for the CVD-COVID-UK/COVID-IMPACT research programme (REC No 20/NE/0161) to access, within secure trusted research environments, unconsented, whole-population, de-identified data from electronic health records collected as part of patients' routine healthcare. The need for informed consent was waived by the North East-Newcastle and North Tyneside 2 research ethics committee. All methods were carried out in accordance with relevant guidelines and regulations.

## References

1. American Psychiatric Association. Diagnostic and statistical manual of mental disorders. 5th ed. Arlington: American Psychiatric Publishing; 2013.

2. Zeidan J, Fombonne E, Scorah J, Ibrahim A, Durkin MS, Saxena S, et al. Global prevalence of autism: A systematic review update. Autism Research. 2022 May 1;15(5):778–90.

3. Salari N, Rasoulpoor S, Rasoulpoor S, Shohaimi S, Jafarpour S, Abdoli N, et al. The global prevalence of autism spectrum disorder: a comprehensive systematic review and meta-analysis. Italian Journal of Pediatrics 2022 48:1. 2022 Jul 8;48(1):1–16.

4. Talantseva OI, Romanova RS, Shurdova EM, Dolgorukova TA, Sologub PS, Titova OS, et al. The global prevalence of autism spectrum disorder: A three-level meta-analysis. Frontiers in Psychiatry. 2023 Feb 9;14:1071181.

5. Bishop-Fitzpatrick L, Rubenstein E. The physical and mental health of middle aged and older adults on the autism spectrum and the impact of intellectual disability. Research in Autism Spectrum Disorders. 2019 Jul 1;63:34–41.

6. Croen LA, Zerbo O, Qian Y, Massolo ML, Rich S, Sidney S, et al. The health status of adults on the autism spectrum. http://dx.doi.org/101177/1362361315577517. 2015 mApr 24;19(7):814–23.

7. NICE | The National Institute for Health and Care Excellence. Overview | Multimorbidity: clinical assessment and management | Guidance | NICE [Internet]. 2016 [cited 2023 Jul 3]. Available from: https://www.nice.org.uk/guidance/ng56

8. Brondino N, Bertoglio F, Forneris F, Faravelli S, Borghesi A, Damiani S, et al. A Pilot Study on Covid and Autism: Prevalence, Clinical Presentation and Vaccine Side Effects. Brain Sciences 2021, Vol 11, Page 860. 2021 Jun 28;11(7):860.

9. Fusar-Poli L, Brondino N, Rocchetti M, Petrosino B, Arillotta D, Damiani S, et al. Prevalence and predictors of psychotropic medication use in adolescents and adults with autism spectrum disorder in Italy: A cross-sectional study. Psychiatry Research. 2019 Jun 1;276:203– 9.

10. May M, Slitzky M, Rostama B, Barlow D, Houseknecht KL. Antipsychotic-induced immune dysfunction: A consideration for COVID-19 risk. Brain, Behavior, & Immunity - Health. 2020 Jul 1;6:100097.

11. Hossain MM, Khan N, Sultana A, Ma P, McKyer ELJ, Ahmed HU, et al. Prevalence of comorbid psychiatric disorders among people with autism spectrum disorder: An umbrella review of systematic reviews and meta-analyses. Vol. 287, Psychiatry Research. 2020.

12. Rydzewska E, Dunn K, Cooper SA. Umbrella systematic review of systematic reviews and meta-analyses on comorbid physical conditions in people with autism spectrum disorder. The British journal of psychiatry : the journal of mental science. 2021 Jan 1;218(1):10–9.

13. Forsyth L, Mcsorley M, Rydzewska E. All-cause and cause-specific mortality in people with autism spectrum disorder: A systematic review. Research in Autism Spectrum Disorders. 2023;105:102165.

14. Tsai SJ, Chang WH, Cheng CM, Liang CS, Bai YM, Hsu JW, et al. All-cause mortality and suicide mortality in autistic individuals: An entire population longitudinal study in Taiwan. Autism. 2023 May 10;

15. Rydzewska E, Hughes-McCormack LA, Gillberg C, Henderson A, MacIntyre C, Rintoul J, et al. General health of adults with autism spectrum disorders – A whole country population cross-sectional study. Research in Autism Spectrum Disorders. 2019 Apr 1;60:59–66.

16. British Medical Association. Recognising the importance of physical health in mental health and intellectual disability: achieving parity of outcomes [Internet]. 2014 [cited 2023 Jul 3]. Available from: https://www.scie-socialcareonline.org.uk/recognising-the-importance-of-physical-health-in-mental-health-and-intellectual-disability-achieving-parity-of-outcomes/r/a11G0000004Gp6QIAS

17. Hirvikoski T, Mittendorfer-Rutz E, Boman M, Larsson H, Lichtenstein P, Bölte S. Premature mortality in autism spectrum disorder. The British Journal of Psychiatry. 2016 Mar 1;208(3):232–8.

18. Catalá-López F, Hutton B, Page MJ, Driver JA, Ridao M, Alonso-Arroyo A, et al. Mortality in Persons With Autism Spectrum Disorder or Attention-Deficit/Hyperactivity Disorder: A Systematic Review and Meta-analysis. JAMA Pediatrics. 2022 Apr 1;176(4):e216401–e216401.

19. Charlson ME, Pompei P, Ales KL, MacKenzie CR. A new method of classifying prognostic comorbidity in longitudinal studies: development and validation. Journal of chronic diseases. 1987;40(5):373–83.

20. Elixhauser A, Steiner C, Harris DR, Coffey RM. Comorbidity Measures for Use with Administrative Data. Medical Care. 1998;36(1):8–27.

21. Van Walraven C, Austin PC, Jennings A, Quan H, Forster AJ. A modification of the Elixhauser comorbidity measures into a point system for hospital death using administrative data. Medical care. 2009 Jun;47(6):626–33.

22. Diederichs C, Berger K, Bartels DB. The Measurement of Multiple Chronic Diseases—A Systematic Review on Existing Multimorbidity Indices. The Journals of Gerontology: Series A. 2011 Mar 1;66A(3):301–11.

23. Stirland LE, González-Saavedra L, Mullin DS, Ritchie CW, Muniz-Terrera G, Russ TC. Measuring multimorbidity beyond counting diseases: systematic review of community and population studies and guide to index choice. BMJ. 2020 Feb 18;368.

24. Quan H, Li B, Couris CM, Fushimi K, Graham P, Hider P, et al. Updating and Validating the Charlson Comorbidity Index and Score for Risk Adjustment in Hospital Discharge Abstracts Using Data From 6 Countries. Am J Epidemiol [Internet]. 2011 Mar 15 [cited 2023 Jul 3];173(6):676–82. Available from: 10.1093/aje/kwq433

25. Hossain MM, Khan N, Sultana A, Ma P, McKyer ELJ, Ahmed HU, et al. Prevalence of comorbid psychiatric disorders among people with autism spectrum disorder: An umbrella review of systematic reviews and meta-analyses. Psychiatry Research. 2020 Mar 18;287:112922– 112922.

26. Rydzewska E, Dunn K, Cooper SA. Umbrella systematic review of systematic reviews and meta-analyses on comorbid physical conditions in people with autism spectrum disorder. The British Journal of Psychiatry. 2021 Jan 1;218(1):10–9.

27. Hwang YI (Jane), Srasuebkul P, Foley KR, Arnold S, Trollor JN. Mortality and cause of death of Australians on the autism spectrum. Autism research : official journal of the International Society for Autism Research. 2019 May 1;12(5):806–15.

28. Nijhof D, Sosenko F, MacKay D, Fleming M, Jani B, Pell J, et al. COVID-19 hospitalisations and mortality in autistic people: A whole-country population study. Manuscript in preparation.

29. Sosenko F, Mackay D, Bhautesh DJ, Fleming M, Pell JP, Hatton C, et al. Understanding covid-19 outcomes among people with Intellectual Disabilities in England. BMC Public Health. 2023;23(2099).

30. Sosenko F, Mackay D, Bhautesh DJ, Fleming M, Pell JP, Hatton C, et al. Understanding covid-19 outcomes among people with Intellectual Disabilities in England. BMC Public Health.

31. Nijhof D, Sosenko F, MacKay D, Fleming M, Jani B, Pell J, et al. COVID-19 hospitalisations and mortality in autistic people: A whole-country population study. Manuscript in preparation.

32. Cassell A, Edwards D, Harshfield A, Rhodes K, Brimicombe J, Payne R, et al. The epidemiology of multimorbidity in primary care: a retrospective cohort study. British Journal of General Practice [Internet]. 2018 Apr 1 [cited 2023 Jul 17];68(669):e245–51. Available from: https://bjgp.org/content/68/669/e245

33. Quan H, Li B, Couris CM, Fushimi K, Graham P, Hider P, et al. Updating and Validating the Charlson Comorbidity Index and Score for Risk Adjustment in Hospital Discharge Abstracts Using Data From 6 Countries. American Journal of Epidemiology. 2011 Mar 15;173(6):676–82.

34. Zhou Y, Chi J, Lv W, Wang Y. Obesity and diabetes as high-risk factors for severe coronavirus disease 2019 (Covid-19). Diabetes/Metabolism Research and Reviews. 2021 Feb 1;37(2):e3377.

35. Saha S, Al-Rifai RH, Saha S. Diabetes prevalence and mortality in COVID-19 patients: a systematic review, meta-analysis, and meta-regression. 2021;

36. Zhang Y, Lin F, Tu W, Zhang J, Choudhry AA, Ahmed O, et al. Thyroid dysfunction may be associated with poor outcomes in patients with COVID-19. Molecular and Cellular Endocrinology. 2021 Feb 2;521:111097.

37. Shmueli G. To Explain or to Predict? https://doi.org/101214/10-STS330. 2010 Aug 1;25(3):289–310.

