## Supplementary Materials for "Making a Case for an Autism-Specific Multimorbidity Index: A Comparative Cohort Study"

*Table 1. List of 35 long-term conditions used as candidates for the ASD-MI.*

| |
|---|
| Alcohol problems |
| Anorexia or bulimia |
| Anxiety & other neurotic |
| Asthma (currently treated) |
| Atrial fibrillation |
| Blindness and low vision |
| Bronchiectasis |
| Cancer - [New]Diagnosis in last five years |
| Coronary heart disease |
| Chronic kidney disease |
| Chronic Liver Disease and Viral Hepatitis |
| COPD |
| Dementia |
| Depression |
| Diabetes |
| Diverticular disease of intestine |
| Epilepsy (currently treated) |
| Heart failure |
| Hearing loss |
| Hypertension |
| Inflammatory bowel disease |
| Irritable bowel syndrome |
| Migraine |
| Multiple sclerosis |
| Peptic Ulcer Disease |
| Parkinson’s disease |
| Prostate disorders |
| Psychoactive substance misuse (not alcohol) |
| Psoriasis or eczema |
| Peripheral vascular disease |
| Rheumatoid arthritis |
| Schizophrenia (and related non-organic psychosis) or bipolar disorder |
| Chronic sinusitis |
| Stroke & transient ischaemic attack |
| Thyroid disorders |

*Table 2. List of the 12 long-term conditions included in the Quan Index.*

| |
|---|
| Myocardial Infarction |
| Congestive Heart Failure |
| Peripheral Vascular Disease |
| Cerebrovascular Disease |
| Hemiplegia or paraplegia |
| Dementia |
| Chronic Pulmonary Disease |
| Rheumatologic Disease |

| |
|---|
| Peptic Ulcer Disease |
| Diabetes without Chronic Complications |
| Diabetes with Chronic Complications |
| Renal Disease |
| Any Malignancy, including Leukemia and Lymphoma |
| Metastatic Solid Tumor |
| Mild Liver Disease |
| Moderate or Severe Liver Disease |
| AIDS/HIV |

*Table 3. Different combinations of the selected LTCs included in the Multimorbidity Index with their respective coefficients and odds ratio.*

| **Predictors** | **Multimorbidity Index** | **Odds ratio** |
|---|---|---|
| None of DIA, CHD, THY | 0 | (base) |
| DIA | 1.21 | 3.35 |
| CHD | 0.71 | 2.03 |
| THY | 0.67 | 1.96 |
| DIA, CHD | 1.92 | 6.82 |
| DIA, THY | 1.88 | 6.55 |
| CHD, THY | 1.38 | 3.97 |
| DIA, CHD, THY | 2.59 | 13.33 |
